# Survey shows limited awareness of tamoxifen-associated uterine cancer risk among breast cancer survivors

**DOI:** 10.64898/2026.02.16.26346375

**Authors:** Yumisa Ellinger, Siddharth Annaldasula, Leonie Stockschläder, Christian Rudlowski, Angela Besserer, Oliver Zivanovic, Christina Kaiser, Tjoung-Won Park-Simon, Jens-Uwe Blohmer, Regine Armann, Kirsten Kübler

## Abstract

**Background:** Tamoxifen is a cornerstone of endocrine treatment for hormone receptor-positive breast cancer, reducing recurrence and breast cancer-specific mortality. However, its use is associated with a small, yet clinically relevant, increase in uterine cancer. As diagnosis of this cancer remains symptom-triggered, it is essential for patients to be aware of this risk and report symptoms promptly for optimal outcomes. We therefore assessed risk awareness among breast cancer survivors while exploring their attitudes towards potential future endometrial surveillance strategies.

**Methods:** Over a 10-month period, a web-based survey was conducted among breast cancer survivors with/without tamoxifen treatment. The mixed-format questionnaire included closed-ended questions and optional free-text comments. Quantitative data were summarized descriptively and analyzed statistically; qualitative responses were reviewed thematically to contextualize survey findings.

**Results:** Of 163 respondents, 154 breast cancer survivors were included in the analysis, 128 of whom had received tamoxifen. Among tamoxifen-associated participants, 60% reported insufficient awareness of the associated uterine cancer risk, and half expressed uncertainty about the adequacy of the current symptom-triggered endometrial evaluation. Despite this, acceptance of tamoxifen therapy was high; only one patient declined treatment over concerns about side effects. Almost all participants (96%) were willing to adopt endometrial surveillance methods, if developed and validated.

**Conclusion:** As evaluation of tamoxifen-associated uterine pathology is symptom-triggered, our data highlight the need for improved and standardized risk communication to promote timely symptom recognition, reporting, and diagnostic evaluation. Moreover, our findings support incorporating patient-reported preferences into the development of future endometrial detection strategies to improve survivorship care.

## Introduction

Tamoxifen, a selective estrogen receptor modulator (SERM), is a well-established adjuvant endocrine therapy for hormone receptor-positive (HR^+^) breast cancer (BC), with a recommended treatment duration of 5 years and, in selected cases, up to 10 years [1]. It is commonly used in premenopausal women with low-to intermediate-risk disease and remains an option for postmenopausal women when aromatase inhibitors are not tolerated [2]. Consistent with this role, meta-analyses of randomized trials in early invasive disease show that 5 years of tamoxifen reduces breast cancer recurrence by up to 47% [3] and breast cancer mortality by about 31% [4]. Moreover, tamoxifen decreases the risk of contralateral breast cancer by approximately 30% in the adjuvant setting [5] and remains a treatment option in the metastatic setting as long as clinical benefit persists [6]. In addition to its therapeutic role, prevention trials in women without BC but at increased risk have demonstrated a reduction in BC incidence of up to 49% with tamoxifen [7, 8]. Reflecting its sustained clinical relevance, tamoxifen is included in the WHO List of Essential Medicines and, even as endocrine options have expanded, remains in use worldwide [9], with recent utilization data indicating overall stability with regional increases [10].

Tamoxifen acts as an estrogen antagonist in breast tissue but has estrogen-agonistic effects on the uterus [11]. Consistent with these stimulatory effects on the endometrium, benign endometrial changes such as polyps and hyperplasia are reported in 8–50% of users [12]. Although uncommon, tamoxifen use is associated with a 2–7-fold increased risk of a secondary cancer in the uterus (tamoxifen-associated uterine cancer, TA-UC) [13]. In current clinical practice, clinical work-up is symptom-triggered with patients counseled to report postmenopausal or atypical bleeding [14], which prompts assessment with transvaginal ultrasound (TVUS) and, when indicated, hysteroscopy with curettage for histological clarification [15].

Despite this well-recognized increase in secondary cancer risk—an exposure profile that might otherwise justify surveillance approaches—current guidelines advise against routine monitoring in asymptomatic tamoxifen users, including TVUS-based approaches [14]. In this screening context, TVUS is limited by low specificity of 64% and low sensitivity of 82% [16], because endometrial thickening is frequent and usually benign [14, 17]. Consequently, TVUS can result in false-positive findings and downstream invasive procedures with associated morbidity, patient anxiety, and healthcare costs [18]. Other monitoring approaches have been evaluated [14], but clinicians still lack an evidence-based surveillance strategy for asymptomatic tamoxifen users, representing a clinically relevant gap in breast cancer survivorship care, especially as TA-UC is associated with more aggressive histological subtypes than sporadic UC [19]. This gap is becoming increasingly relevant as more women live longer after BC [20], deepening the clinical relevance of treatment side effects. In addition, the rising incidence of early-onset BC [21] further amplifies this issue by expanding the premenopausal population for whom tamoxifen is frequently used. They also have an increased risk of TA-UC [22], and as survivorship often intersects with fertility planning and pregnancy-related considerations in this group, timely detection and management of uterine pathology is of high importance.

Survivorship follow-up is increasingly shifting toward individualized, risk-adapted strategies [20, 23]. This development may be particularly relevant for tamoxifen-treated patients, for whom a substantial clinical benefit in BC outcomes coexists with a small, yet clinically relevant, risk of secondary UC that might be mitigated through earlier detection. We thus assessed BC survivors’ willingness to consider endometrial surveillance to understand acceptability of future approaches should they become available. Further, until such evidence-based strategy is validated, management remains symptom-triggered and thus depends on patients recognizing and reporting symptoms. We therefore also assessed patients’ awareness of TA-UC risk. Together, understanding patient perspectives is central to facilitate individualized survivorship care and to guide implementation of future risk-based follow-up strategies.

## Material and methods

### Study design, participants, and survey instrument

From May 23, 2024, to March 31, 2025, an open, web-based voluntary survey was conducted in Germany among patients with a history of BC and current or prior exposure to antihormonal treatment. Inclusion criteria were: (i) a diagnosis of BC; (2) age ≥18 years; and (3) the ability to provide informed consent. To facilitate broad participation, convenience sampling was applied to recruit participants; as no complete list of eligible individuals (sampling frame) was available, calculation of a response rate was not possible.

The questionnaire was designed by the study team (Y.E., R.A., K.K.). The survey (**Supplementary File S1**) was administered using LimeSurvey, with participant access provided via the fragdiepatienten.de portal, a platform operated by the *Deutsches Krebsforschungszentrum* (DKFZ) [24]. The survey link and QR code were distributed through flyers and email communications to clinics and physician offices across Germany, with a request to forward them to eligible patients; additional dissemination occurred via patient networks and advocacy groups.

A mixed-methods design was used to collect quantitative and qualitative data. The self-administered survey contained 32 items in total; due to the branching logic that directed participants to different follow-up questions, each participant answered a maximum of 29 questions. The survey included closed-ended single- and multiple-choice questions as well as optional open-ended free-text fields. All closed-ended items were mandatory, and most provided an “I do not know” response option to capture uncertainty or lack of knowledge.

Key domains assessed included: (i) perceived sufficiency of information regarding TA-UC risk; (ii) perceived safety of symptom-triggered monitoring; (iii) willingness to adopt new endometrial monitoring methods if available; and (iv) whether TA-UC concerns influenced treatment refusal. Baseline characteristics collected included age and BC treatment history.

### Ethics statement

Prior to participating, respondents were presented with an informed consent statement outlining the study purpose, the anonymous nature of data collection and analysis, and the possibility of publication. Informed consent was obtained electronically from all participants. Data processing was conducted in accordance with Art. 6(1)(a) and Art. 9(1) for health data of the EU’s General Data Protection Regulation (GDPR). According to §15(1) of the professional code of conduct of the local Medical Association, formal ethics committee approval was not required for anonymously collected survey data. The study was conducted in compliance with national law and in accordance with the 1964 Declaration of Helsinki and its later amendments.

### Data analysis and statistics

Quantitative responses were aggregated, analyzed, and visualized using R (version 4.4.1) in Rstudio, employing the *tidyverse, dyplr, ggplot2*, and *ComplexUpset* packages. Results are reported as counts and percentages. A two-tailed Fisher’s exact test was used to compare frequencies of categorical answers between subgroups; within groups, binomial tests were used to assess whether the distribution of binary answers (yes/no) deviated from an equal distribution (50/50 split) under the null hypothesis of equal probability for each response option. Statistical significance was set at *p*<0.05. Error bars represent the standard error (SE) of the proportion, expressed as percentage and calculated as SE = √(*p* × (*1* − *p*) /*n*) × *100*, where *p* denotes the observed proportion and *n* the sample size. Qualitative free-text responses were reviewed thematically to identify recurring patterns and illustrative themes.

## Results

### Patient characteristics

A total of 163 participants completed the survey (**Figure 1**); nine individuals without a history of BC were excluded, resulting in a final sample of 154 patients. Most participants (96%) were diagnosed and treated in Germany, providing a highly homogenous cohort in terms of the healthcare setting (**Table 1**). Based on responses to the initial tamoxifen therapy question (yes/no), participants were divided into two subgroups: 128 patients treated with tamoxifen (study cohort) and 26 without tamoxifen exposure (controls). Of the study cohort, 73 patients (57%) were receiving tamoxifen; most (n=70) were in the initial treatment phase. Almost all participants (>90%) had been diagnosed with HR^+^ BC >1 year before the survey; hence, our sample is mostly composed of survivors and patients further along in their treatment journey. The estimated median age fell within the 46–65-years range. In the tamoxifen-treated cohort, 97 patients (76%) had no endometrial changes and 31 (24%) did. Among those with abnormalities, 29 (94%) were benign; one was indeterminate, and one was atypical endometrial hyperplasia, a precancerous lesion associated with an increased risk of UC. No cases of TA-UC were reported.

**Table 1.** Baseline clinical characteristics of participants with prior breast cancer (BC).

| Variable | Category | Number (%) |
| --- | --- | --- |
| Age at survey (n=154) | ≤45 years | 33 (21.4) |
|  | 46–65 years | 113 (73.4) |
|  | 66–75 years | 8 (5.2) |
|  | ≥76 years | 0 (0.0) |
| BC diagnosis-to-survey interval (n=154) | ≤1 year | 15 (9.7) |
|  | 1–5 years | 75 (48.7) |
|  | ≥5 years | 64 (41.6) |
| Disease status & treatment (n=154) | Treatment not yet started | 0 (0.0) |
|  | Adjuvant treatment* ongoing, no tamoxifen | 13 (8.4) |
|  | Adjuvant treatment ongoing, tamoxifen ongoing | 70 (45.5) |
|  | Adjuvant treatment ongoing, tamoxifen completed** | 5 (3.2) |
|  | Adjuvant treatment completed, no tamoxifen | 11 (7.1) |
|  | Adjuvant treatment completed, tamoxifen completed | 33 (21.4) |
|  | Relapse, no tamoxifen | 1 (0.7) |
|  | Relapse, tamoxifen ongoing | 3 (1.9) |
|  | Relapse, tamoxifen completed | 6 (3.9) |
|  | Advanced disease/palliative, tamoxifen completed | 1 (0.7) |
|  | Unknown disease status, no tamoxifen | 1 (0.7) |
|  | Unknown disease status, tamoxifen completed | 10 (6.5) |
| Country of diagnosis & treatment (n=154) | Diagnosis & treatment in Germany | 147 (95.5) |
|  | Diagnosis in Germany, treatment abroad | 1 (0.6) |
|  | Diagnosis abroad, treatment in Germany | 0 (0.0) |
|  | Diagnosis & treatment abroad | 6 (3.9) |
| History of tamoxifen intake (n=154) | No tamoxifen | 26 (16.9) |
|  | Tamoxifen users | 128 (83.1) |
|  | Completed treatment | 45 (35.2) |
|  | Current treatment | 83 (64.8) |
| **Duration of tamoxifen intake (n=128)** | ≤1 year | 21 (16.4) |
|  | 1–3 years | 54 (42.2) |
|  | 4–5 years | 22 (17.2) |
|  | ≥5 years | 31 (24.2) |
|  | Unknown | 0 (0.0) |

\*, adjuvant treatment indicates primary disease status and may include surgery, chemotherapy, radiotherapy, and antihormonal treatment (tamoxifen, aromatase inhibitors, or gonadotropin-releasing hormone [GnRH] analogues; \*\*, may include sequential/switch therapy or discontinuation).

**Figure 1.**
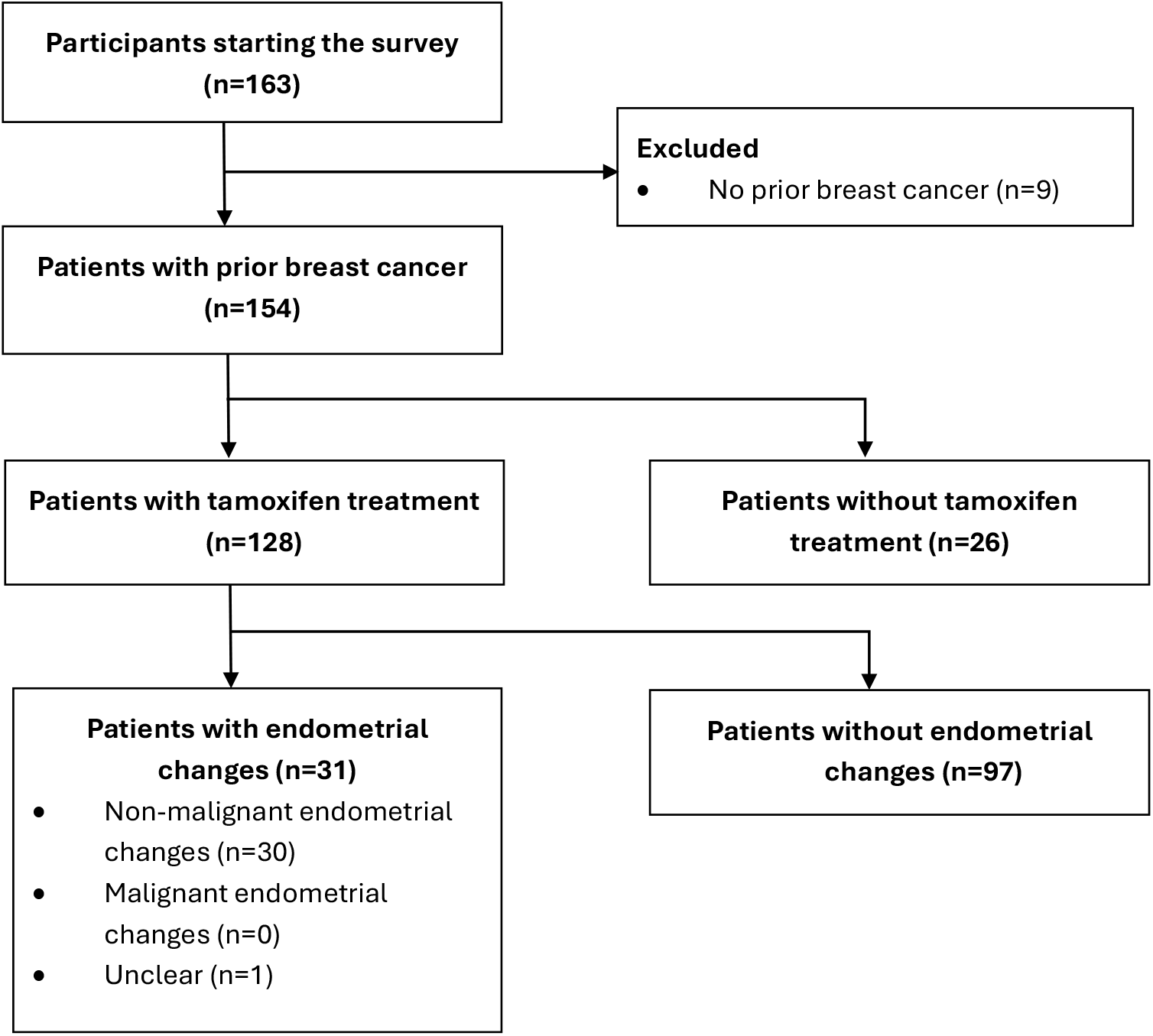
Allocation of patients. CONSORT flow diagram depicting the inclusion and exclusion of participants in the survey. Non-malignant endometrial changes include both benign and precancerous findings.

### Patient awareness of TA-UC risk

We first assessed whether participants felt sufficiently informed about the risk of TA-UC. Among the 128 individuals with a history of tamoxifen use, only 46 (35.9%) reported sufficient awareness, while 77 (60.2%) reported insufficient awareness, a proportion significantly higher than expected by chance (*p*=0.007; binomial test; **Fig. 2a**). In the control cohort without tamoxifen exposure, awareness was low as expected (5/26, 19.2%). Notably, awareness among tamoxifen users was not higher than in controls (*p*=0.1; Fisher’s exact test), suggesting that tamoxifen treatment and accompanying counseling did not substantially improve risk knowledge beyond baseline awareness.

**Figure 2.**
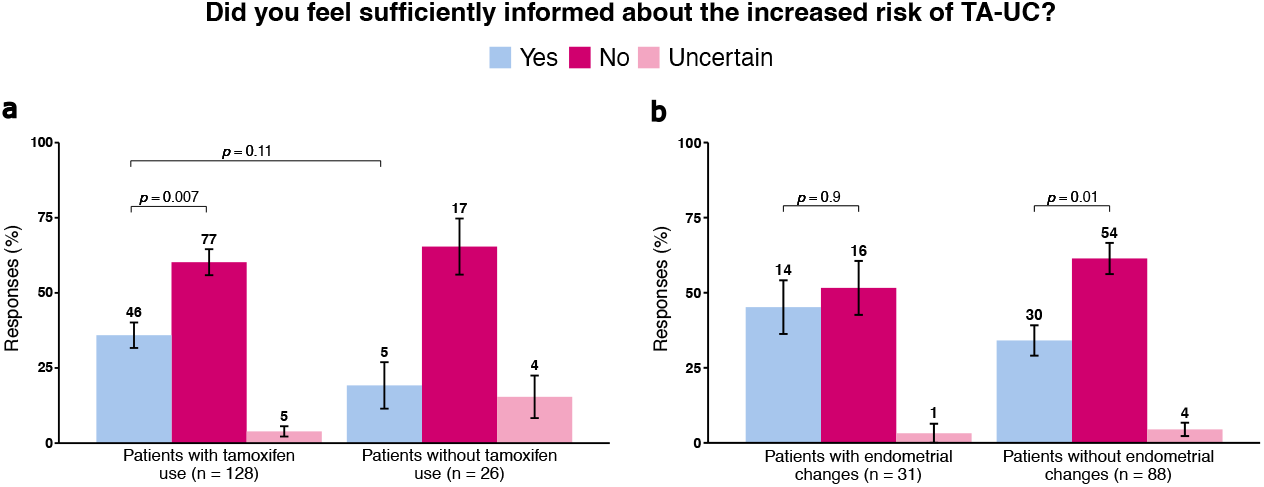
Awareness of risk. Bar plots showing response frequencies regarding tamoxifen-associated uterine cancer (TA-UC) risk. Error bars reflect the standard error of the proportion (based on the binomial distribution); numbers above bars indicate counts per group. **(a)** Responses from all patients, stratified by tamoxifen use; significance analyses by binomial test and two-sided Fisher’s exact test. **(b)** Comparison among tamoxifen-treated patients with and without endometrial changes; significance analyses by binomial test.

In deviation from guideline recommendations, 71% of the tamoxifen cohort reported undergoing TVUS without symptoms. Only 10 patients (7.8%) reported TVUS performed for symptoms as recommended. To explore the effect of TVUS and associated physician–patient communication opportunities on awareness, we stratified the tamoxifen-treated cohort based on whether patients had undergone an examination (91 yes vs. 37 no). Patients who underwent only sporadic or no TVUS were more likely to report insufficient information (31/37; 83.8%) than patients who underwent repeated examinations (46/91; 50.5%; *p*=0.0006; Fisher’s exact test). When stratified by diagnosis of endometrial pathology, unaffected patients were predominantly uninformed about the risk of TA-UC (61% vs 34%; *p*=0.01; binomial test; **Fig. 2b**), whereas in affected patients the diagnosis reduced the information gap, with unawareness and awareness reported at similar rates (52% vs 45%, *p*=0.9; binomial test). Thus, while half of the patients undergoing examinations still reported insufficient awareness, suggesting that physician contact does not guarantee adequate education, the diagnosis of endometrial pathology appeared to narrow the awareness gap but did not eliminate the overall information gap.

In the control cohort of 26 patients, tamoxifen was offered to five women as adjuvant therapy, bringing the total number of women with a tamoxifen recommendation across the study to 133. Of these five, two opted for aromatase inhibitors for undisclosed reasons, one refused because of a potential interaction with her antidepressant drug, and one, due to a *BRCA1* germline mutation requiring oophorectomy, was advised to take an aromatase inhibitor. Three of the five felt sufficiently informed about the risk of TA-UC, with one ultimately refusing tamoxifen due to concerns about side effects, possibly including TA-UC. Thus, overall, only one patient (1/133; 0.8%) reported declining tamoxifen, suggesting that treatment refusal was uncommon in this cohort, although persistent knowledge gaps regarding TA-UC risk may have influenced such decisions.

### Sources of information on TA-UC risk

Next, we examined from which sources patients obtained information about TA-UC risk. Overall, 47.7% of patients with a history of tamoxifen treatment reported receiving information from their physician, making this the most cited source (**Fig. 3**). A similar proportion reported relying on online sources (41.4%). The response option “other sources” was selected by 15.6% and included the questionnaire itself, as illustrated by three patients’ free-text comments stating, “first informed via this survey”. Similarly, in the control group of patients without a history of tamoxifen use, physician consultation and online sources were equally common (both 42.3%). These observations suggest that TA-UC awareness may be influenced less by the type of information source and more by gaps in physician-provided counseling.

**Figure 3.**
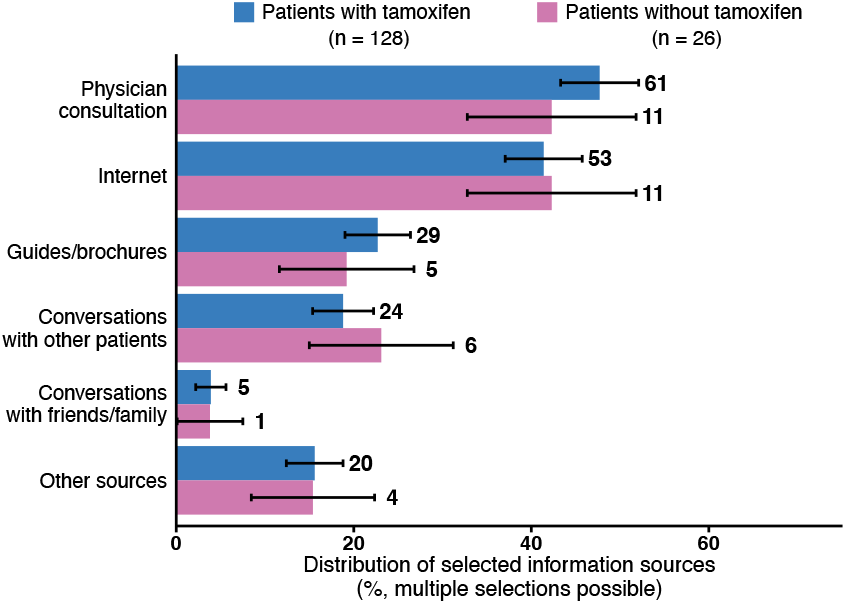
Information sources on risk. Bar plots depicting the distribution of information sources, stratified by use of tamoxifen. Total percentages may exceed 100% as multiple responses were allowed. Error bars reflect the standard error of the proportion (based on the binomial distribution); numbers above bars indicate counts per group.

### Patient perception of safety

Of the patients with a history of tamoxifen treatment, half (64/128, 50%) reported a subjective sense of safety regarding TA-UC, while the remainder felt unsafe (41/128, 32%) or were unsure (23/128, 18%). Reassurance was more often reported by those undergoing TVUS (60.4% vs. 24.3%, *p*=0.0004; Fisher’s exact test), even though TVUS is not a reliable or recommended method of surveillance. Together with free-text responses, in which 15 patients requested closer monitoring, these findings show that the current approach fosters a false sense of reassurance, while also highlighting patients’ unmet need for a genuine sense of safety. When stratified by the presence/absence of endometrial alterations, perceived safety did not differ significantly between groups (both *p*>0.05; Fisher’s exact test). However, patients with endometrial changes were less often uncertain about their safety (1/31; 3.2%) than those without (20/88; 22.7%; *p*=0.01; Fisher’s exact test; **Fig. 4**), indicating that the detection of an abnormality compelled patients to form a clearer opinion.

**Figure 4.**
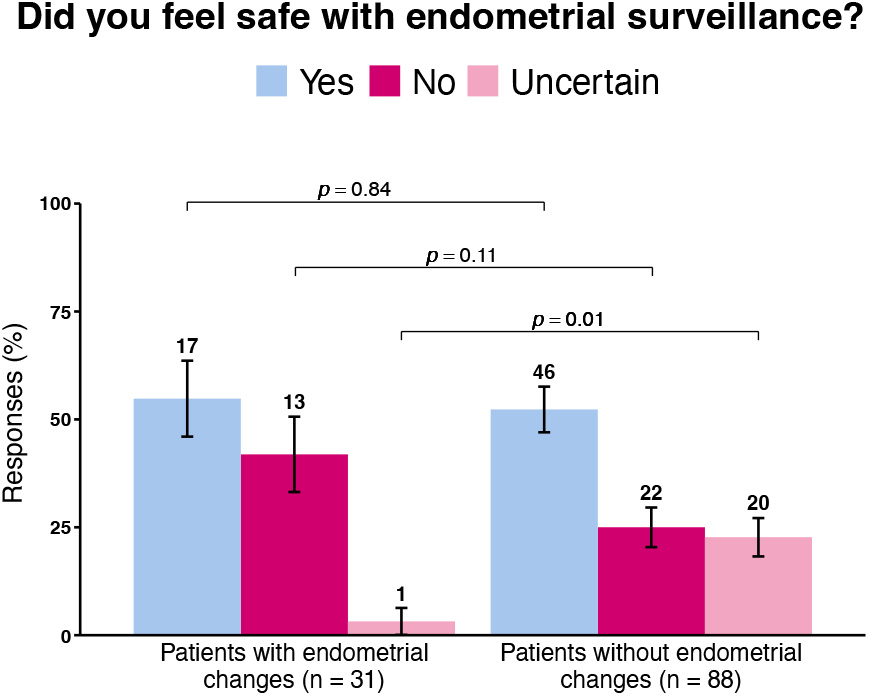
Perceived subjective safety of endometrial surveillance. Bar plots depicting the distribution of responses among tamoxifen-treated patients, comparing between patients with and without endometrial changes. Error bars reflect the standard error of the proportion (based on the binomial distribution); numbers above bars indicate counts per group; significance analysis by two-sided Fisher’s exact test.

### Openness to potential future options for endometrial surveillance

In a hypothetical scenario, patients were asked whether they would consider using endometrial surveillance methods if such options became available and were proven effective and safe. Across the tamoxifen-treated group, 123 patients (96%) expressed such willingness. This openness was similarly high among patients without endometrial alterations (85/88; 96.6%) and those with such changes (29/31; 93.5%; **Fig. 5a**). Subgroup analyses showed consistently high willingness, irrespective of whether participants reported prior uterine assessments (e.g., TVUS or other methods described as MRI, CT, or smears in free-text responses; 93/97; 95.9%), felt safe (59/64; 92.2%), or recalled prior TA-UC risk communication (44/46; 95.7%; **Fig. 5b**). Even among those who reported having prior assessments and who felt both safe and sufficiently informed about TA-UC, most (29/31; 93.5%) still expressed interest in adopting non-invasive surveillance approaches, if developed and validated. Only one patient (0.8%) rejected this idea, while four (3.3%) were unsure. Although the questionnaire did not capture health insurance type, many respondents (53.9%) indicated they would consider personal payment for future monitoring methods. This willingness did not differ between patients with endometrial alterations (19/31; 61.3%) and patients without (44/88; 50%; *p*=0.3, Fisher’s exact test; **Fig. 5a**). Together, these findings highlight substantial patient interest in future surveillance approaches and even readiness to self-fund them, underscoring the importance of establishing and validating effective new methods.

**Figure 5.**
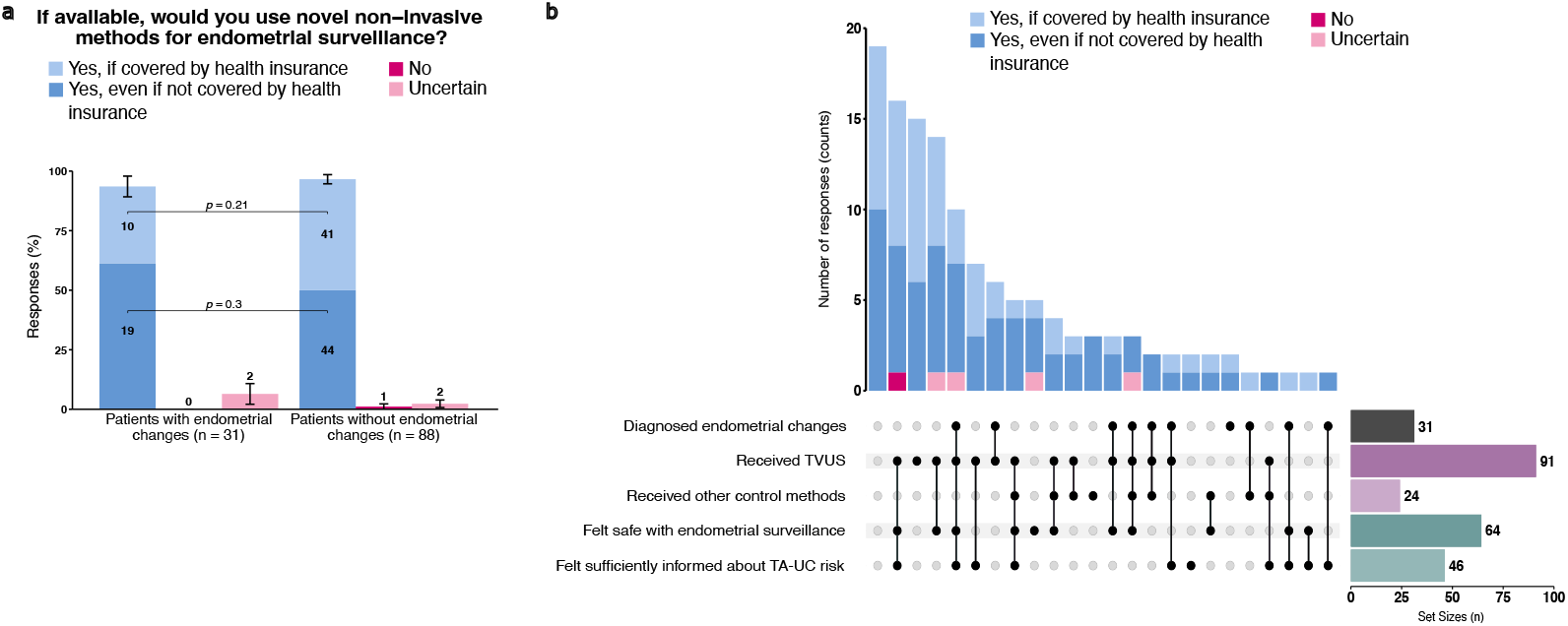
Acceptance of new endometrial surveillance methods when taking tamoxifen. **(a)** Stacked bar plot showing responses to whether patients would make use of novel endometrial monitoring methods, when available, stratified by presence or absence of endometrial alterations. Error bars reflect the standard error of the proportion (based on the binomial distribution) for all answers in a group; numbers in bars indicate counts per subgroup; significance analyses by two-sided Fisher’s exact test. **(b)** Upset plot illustrating the overlap of patient-reported experiences regarding endometrial alterations, tamoxifen-associated uterine cancer (TA-UC) risk communication, perceived subjective safety, and type of endometrial surveillance (transvaginal ultrasound [TVUS] or other examinations). Bars above the intersection matrix represent the number of patients reporting each specific combination; colors indicate patient willingness to use novel non-invasive surveillance methods if they become available. Bars to the right show the total number of patients reporting each category.

## Discussion

This survey revealed low patient awareness of TA-UC risk, which is concerning given that current management relies on symptom-prompted evaluation. At the same time, respondents expressed strong willingness to consider future surveillance approaches, should they become available. Together, these findings highlight a need for improved and standardized risk communication and support the development of individualized, risk-adapted follow-up strategies.

With 96% of participants diagnosed and treated within the same healthcare system, the cohort is highly homogeneous, strengthening the reliability of our findings by minimizing variability in care and reducing confounding factors. The majority of participants were younger than both the median age at UC diagnosis in Germany (67 years [25]) and the typical age range reported for TA-UC (55–69 years [26]), which may partly explain the absence of TA-UC in our cohort. Nevertheless, 24% of patients reported endometrial changes, in line with previously reported rates of 8–50% in primarily postmenopausal patients [12]. In addition, the fact that most patients had been in treatment for an extended period suggests that they can offer a longer-term perspective on TA-UC risk communication, strengthening confidence in the interpretability of our findings.

Although the association between tamoxifen and UC has been recognized since the 1980s [27-29] and been the subject of decades of research and physician education [30], literature on BC patients’ knowledge regarding TA-UC remains scarce. In a 2015 survey of 299 patients in South Korea, 57.9% of the participating BC patients reported awareness of the increased risk [31]. In a 2020 exploratory study on 408 patients in the UK assessing TA-UC awareness among women at higher risk for BC considering preventive treatment, only 26.2% were informed about TA-UC as a potential harm [32]. Our finding that 35.9% of patients felt well-informed is broadly consistent with these previous findings, falling between the two estimates. This general alignment supports the robustness of the observed association, with variations potentially reflecting differences in methodological approaches (e.g., assessment of factual knowledge vs. perceived sufficiency of information) or structural differences between national healthcare systems and counseling practices.

Whether due to the unlikely omission by physicians or to patients’ non-internalization of information under stress or cognitive overload, our findings reveal a gap in effective patient counseling. Addressing this gap may require standardized, repeated risk communication without undermining tamoxifen acceptance or adherence. When patients remain uncertain about their individual risk and the adequacy of symptom-triggered endometrial evaluation, this uncertainty may fuel reliance on non-recommended measures such as TVUS—not for their clinical value, but as reassurance—consistent with evidence that the impulse to reassure is a documented driver of unnecessary testing [33]. Despite clear evidence that routine TVUS in asymptomatic tamoxifen users confers no benefit and may even be harmful [34], our results show a substantial number of patients undergoing TVUS. This likely reflects a combination of patient requests for reassurance, the absence of validated surveillance strategies [34] and—within the German setting—TVUS being readily available as an individual health service (IGeL).

To our knowledge, this is the first study to assess patients’ willingness to consider endometrial surveillance, and despite the hypothetical nature of the scenario, respondents expressed broad openness. Moreover, although the German healthcare system provides broad public coverage and fosters the expectation that most necessary care is already covered [35], many patients still expressed willingness to self-fund future monitoring methods, further underlining the importance of this issue from the patient perspective. Together these findings support the need for effective, individualized, risk-adapted strategies, which should be precise and patient-friendly, to offer genuine reassurance to asymptomatic patients without exposing them to unnecessary interventions.

Our survey was intentionally short, not based on standardized questionnaires, and relied on self-reported data, all of which are limitations and restricted our ability to fully contextualize patients’ responses. In addition, recruitment based on convenience (rather than random sampling) limits generalizability, as does our targeting of only German residents, whose healthcare system provides broad public coverage (which does not necessarily apply to other healthcare structures). Furthermore, the web-based format likely introduced a selection bias towards younger, more digitally literate patients, consistent with most participants being younger than the median age at BC diagnosis in Germany (65 years [36]). Finally, although the survey was directed at all BC patients, mentioning tamoxifen in the title may have disproportionately attracted tamoxifen users, resulting in a smaller control group and limiting our ability to assess how awareness of TA-UC risk might influence adjuvant treatment choices.

Together, our findings underline the need for tailored, clear information to minimize uncertainty, as patient awareness appeared to depend primarily on communication from healthcare providers rather than individual initiative. Of note, some patients listed MRI, CT, and Pap smears as currently employed “other methods”, suggesting that endometrial surveillance is at times confused with cervical cancer screening, reinforcing the importance of clear counselling. While physicians may worry that detailing severe side effects of tamoxifen could undermine adherence to tamoxifen, an indispensable pillar in HR^+^ BC treatment, transparent harm–benefit discussions are essential for informed decision-making and especially important when management relies on symptom-triggered evaluation. Our study highlights the need for further research into new, personalized, and preferably non-invasive follow-up strategies— such as biomarker-based approaches or digital health tools—that offer improved diagnostic specificity. This is now becoming feasible, as a recent study has, for the first time, identified the specific mechanism by which tamoxifen increases the risk of UC [37]. Such advances could enable effective prevention and shift the dialogue: rather than reducing concerns about severe side effects through omission, transparent and stratified risk communication combined with precise, individualized follow-up care can built trust and promote truly patient-centered survivorship medicine.

## Conclusion

This pilot study demonstrates that HR^+^ BC patients taking tamoxifen perceive gaps in information about TA-UC risk and, when informed, express strong support of the adoption of personalized surveillance methods to ensure safety. These findings emphasize the value of integrating patient voices into tamoxifen-related care to guide future research priorities and inform the development of new clinical follow-up strategies.

## CRediT authorship contribution statement

**Yumisa Ellinger:** Data curation, Formal analysis, Investigation, Visualization, Writing – original draft. **Siddharth Annaldasula**: Formal analysis, Writing – review & editing. **Leonie Stockschläder**: Resources, Writing – review & editing. **Christian Rudlowski**: Resources, Writing – review & editing. **Angela Besserer**: Resources, Writing – review & editing. **Oliver Zivanovic**: Resources, Writing – review & editing. **Christina Kaiser**: Resources, Writing – review & editing. **Tjoung-Won Park-Simon**: Resources, Writing – review & editing. **Jens-Uwe Blohmer**: Resources, Writing – review & editing. **Regine Armann**: Methodology, Project administration, Writing – review & editing. **Kirsten Kübler**: Conceptualization, Supervision, Methodology, Funding acquisition, Writing – review & editing.

## Data availability

De-identified survey data (closed-ended items) and the codebook will be deposited in Mendeley Data and made publicly available upon publication.

## Declaration of AI-assisted technologies

During the preparation of this work authors used generative AI for programming support and language editing; all content was verified by the authors.

## Acknowledgements

We thank all patients for participating in this survey. We are grateful to fragdiepatienten.de for helping implement the survey and PINK!, Frauenselbsthilfe Krebs Bundesverband, yeswecan!cer and Aktion Pink for sharing it. We are indebted to Cindy Körner, Barbara Brückner, Ulla Ohlms, and Esther Wiedemann for help disseminating it.

